# A systematic literature review to evaluate the effect of maternal education on under-five mortalities in Nigeria, with recommendations for sustainable development

**DOI:** 10.1101/2024.07.18.24310653

**Authors:** Dominic D. Umoru

**Affiliations:** Department of Paediatrics, Maitama District Hospital

**Keywords:** maternal, education, under-five, mortality, Nigeria

## Abstract

**Introduction:** *The* second target of the SDG3 hopes to reduce under-5 mortalities (U5M) to at least as low as 25 per 1000 live births by 2030. However, U5M remain high in Nigeria and maternal education has been recognized as a predicting factor. This systematic literature review aims to evaluate the relationship between maternal education and under-five mortalities in Nigeria and make recommendations.

**Methods:** A systematic search of PubMed, CINAHL, EMBASE and Maternity and Infant Care databases was conducted between 1st August 2021 and 31^st^ July 2022. Included articles were appraised using Critical Appraisal Tool for Analytical Cross-sectional Studies developed by the Joanna Briggs Institute (JBI), 2017. The Preferred Reporting Items for Systematic Review and Meta-Analyses (PRISMA, 2009) guidelines was used for data extraction.

**Result:** A total of 215 articles were retrieved out of which 33 were removed due to duplications. After screening, 27 were assessed out of which 14 were finally included. The result indicates that there was less odd of U5M among children whose mothers had at least primary education and suggests that the risk of dying further reduces with higher levels of maternal education. This result corroborates that of earlier studies.

**Conclusion:** Reduction in U5M depends on understanding of the impact of maternal education. Therefore, policies should strive to bridge the gaps in female literacy by creating better access to education for Nigerian women.

**Key message:** U5M remains high in Nigeria despite an improving global figure. Aside the direct causes, maternal education has been identified as one remote predictor of child deaths before the age of 5. Although some isolated studies have tried to prove this, there is still need for more evidence to support this thinking, given the potential of such findings to influence policies related to child health. Therefore, this SLR hopes to synthesize evidence based on what is already known with the goal to influence local policies on child survival.

## Introduction

The United Nation’s Sustainable Development Goals (SDGs) to address the limitations observed in the Millennium Development Goals (MDGs) and to chart the path for a new global development ^1^. It therefore hopes to improve on the gains of the MDGs and to formulate a road map for a more sustainable development. The 17 goals and 169 targets are also meant to build upon and extend the achievements of the MDGs by integrating the dimensions of sustainable development ^2^. Virtually all the SDGs may be viewed as being health-related either directly or indirectly. However, SDG3 centers tangentially on health and its second target hopes to among other things reduce under-5 mortalities (U5M) to at least as low as 25 per 1000 live births by 2030.

According some experts^3^, under-5 mortality rate (U5MR) among other things, reflects the availability of maternal and child health services, family income, as well as the over-all safety of the child’s environment. Additionally, child mortality is a fundamental indicator of the health systems performance in countries^4^. Wide differences exist in the U5MRs between high- and low-income countries with countries with the United Kingdome having an U5MR of 4.2/1000 live births in 2020^5^ and Nigeria, a rate of 113.8/1000 live births^6^. Although there has been a progressive decline since the 1990s, the current figures are still unacceptably high when compared with the global target of 25/1000 live births by 2030.

According to Sharma^7^, although biomedical interventions can orchestrate short-term health deliverables, only political and social interventions can engineer durable and holistic results. Such variables have been grouped into three major categories: community, household, and individual factors^8^. Therefore, beyond direct causes of death, several socio-economic factors contribute to U5M and many reports from across the world are congruent on the role played by maternal education. For example, a decomposition analysis conducted in 32 sub-Saharan countries over a period of 16 years indicated that among four major risk factors for U5M, poor maternal education was frequent in more countries than any other factor studied^8^. In a related Ethiopian case control study, a significantly lower odd of dying was reported among under-five children born to mothers who had at least primary literacy^9^. Similarly, a cross-sectional study of 3,975 women in Ghana noted low maternal education as a significant correlate of U5M^10^. Back in Nigeria, a descriptive study based on the 2013 Demographic and Health Survey (DHS) reported sub-national discrepancies in U5M, and the results demonstrated that a 72.5% of deceased children in the north were born to mothers without basic education^11^. Similarly, in another Nigerian study^12^, basic primary literacy among mothers was associated with low rates of under-five deaths, further buttressing the pertinence of the role played by maternal education on child survival.

Aside general maternal education, other reports have debuted the significance of the grade of mother’s education to U5M. For example, it has been established that the level of maternal education may be the real determining factor rather than absolute ability to read or write, or the otherwise^9^. Again, these connections between U5M and maternal education underscores the interdependence of the different SDGs and the need for a collaborative and cross-cutting approach to health interventions. Based on available data^6^, U5M is unacceptably high in Nigeria when compared with the global targets, and poor maternal education is an identifiable gap that needs to be addressed to mitigate the menace and move towards achieving the global targets in the country. Although some studies have highlighted this, there is dearth of systematic reviews aimed specifically to elucidate the effect of the former on the latter in the country, further justifying the need for this study, more so that the writer did not find any in his search. This systematic literature review (SLR) therefore hopes to answer the question “Is there an association between maternal education and under-five mortalities in Nigeria?” by synthesizing evidence from available literatures. The review also hopes to determine how gaps in maternal education could impact the attainment of the SDG target for U5MR and the SDG3 in general. The PICO (Population, Intervention, Comparison and Outcome) format was used to develop the research question. In this process, Population = Nigerian Mothers; Intervention = maternal education; Comparison = Illiteracy; Outcome = Under-5 deaths.

## Methodology

### Search Strategy

A thorough literature search on “the role of maternal education on Under-Five Mortalities in Nigeria” was undertaken. A systematic search was done using PubMed, CINAHL, EMBASE, and Maternity and Infant Care. In addition, other electronic databases such as African Journal Online (AJOL), Google scholar, Scopus, clinical websites, and references were also searched. The search terms were pre-defined, and the key words included “maternal” “mother” “Nigeria” “education” “literacy” “illiteracy” “child” “children” “under-five” “mortality” “fatality” and “death”. The search terms included “maternal” “mother*” “literacy” “literate” “illiteracy” “child*” “under-5” “under-five” “mortalit*” “death*” “fatalit*”, and “Nigeria” was used as a filter. In addition, other search filters included year “2002 to 2022” (20 years) and publications in “English” language. Likewise, the Boolean operators, “AND” was used, for example, to search for “maternal AND “education”, while “OR” was used, for example, to search for “education” OR “awareness”. The search strategy was developed in the PubMed database and then extended to other electronic databases.

### Study selection

The SLR was formulated and reported using the Preferred Reporting Instrument for Systematic reviews and Meta-analysis (PRISMA) checklist^13^. The duplicated studies were excluded while the inclusion and exclusion criteria were then used to select the studies to be included.

### Inclusion criteria

Peer-reviewed journal publications with studies carried out in Nigeria including cross-sectional, cohort and case-control studies assessing the effect of maternal education (independent variable) on U5M (outcome variable) as main, or part of study objective were included. U5M in this study was defined as all child deaths occurring after birth up till before the fifth birthday (birth till 59 months). All such human studies conducted from August 1^st^ 2002 to July 31^st^ 2022 were included.

### Exclusion criteria

All similar studies with neonatal mortalities, NNM (birth to 28 days), post-neonatal mortalities, PNNM (after 28 days till 59 months), infant mortalities, IM (birth till 1 year) and child mortalities, CM (after 1 year till 59 months) as main outcome variable, but not including U5M as outcome variable were excluded. Similarly, studies without clearly stratified maternal educational level were excluded. All papers published before 1^st^ August 2002 were also excluded.

### Data extraction

Data was extracted from full texts of included studies and recorded using a table (Table 1). The data of interest from the selected studies included study type (cross sectional survey, cohort or case-control), sample size, mothers’ educational status (including stratification) and the study setting (whether it was national or sub-national). Also of interest was the description of the independent and outcome variable, and the description of evidence of association. The extraction was based on the PRISMA guidelines^13^, displayed in Figure 1. The publication title, objectives, and methodology of each of the study were used to determine whether they were relevant to the review or not. The first step was to exclude articles based on their titles, including those that did not match the goals of the current review. The whole content of the remaining articles was then assessed in detail and selected based on their focus on the intervention (maternal education) and the outcome, being U5M. To these regards, studies with sample sizes considered as being too small for their methodologies were excluded. Likewise, only studies that were cross-sectional in nature, case-control and cohort reports were included. The independent variable stratification was based on; no formal education (none), primary, secondary and higher education while the outcome variable was under-five mortality. All studies that met the above criteria were included. To display and describe included studies, table used by Kiross et al^14^ in a similar SLR was adapted (Table 1).

**Table 1:**
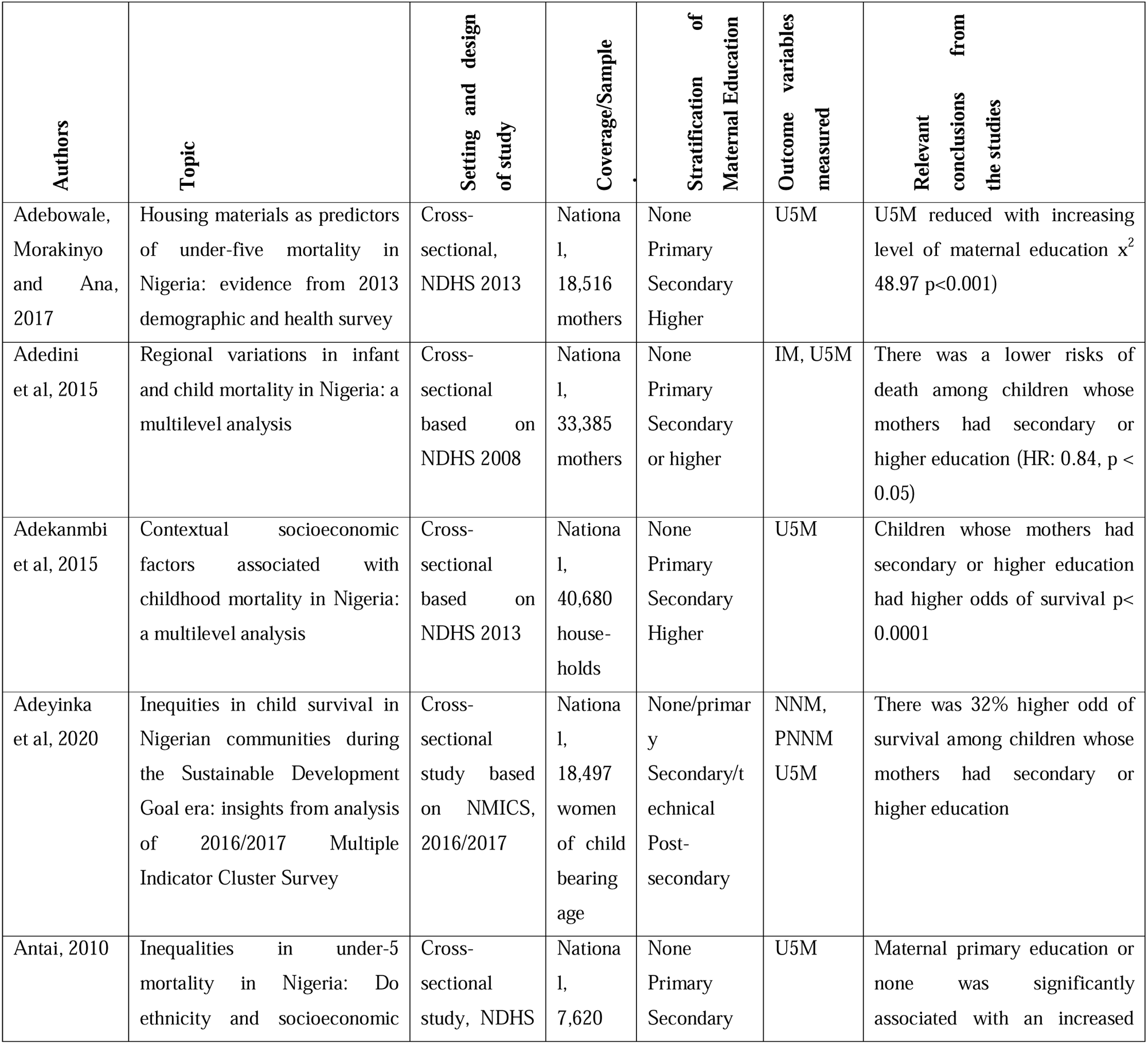
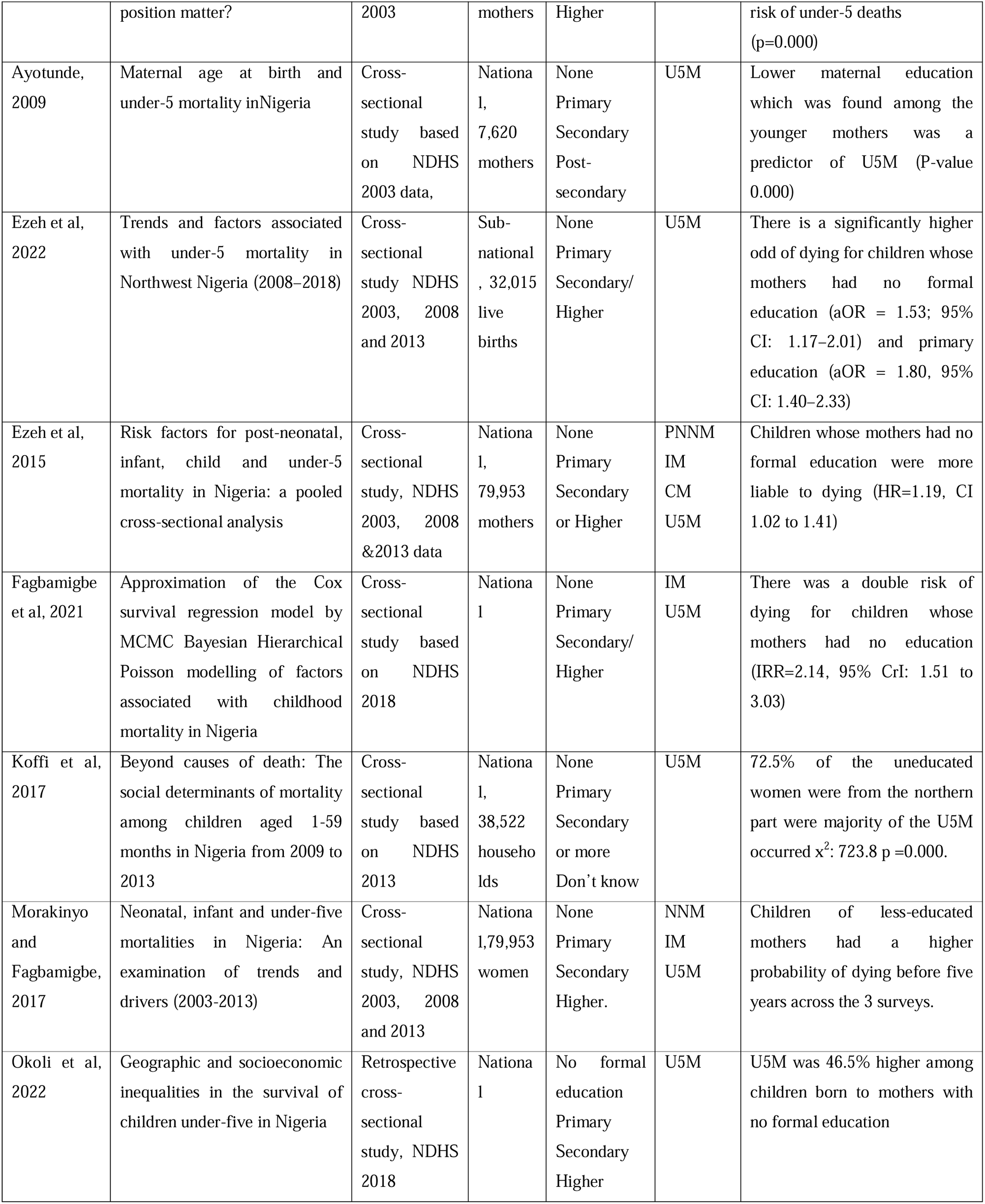
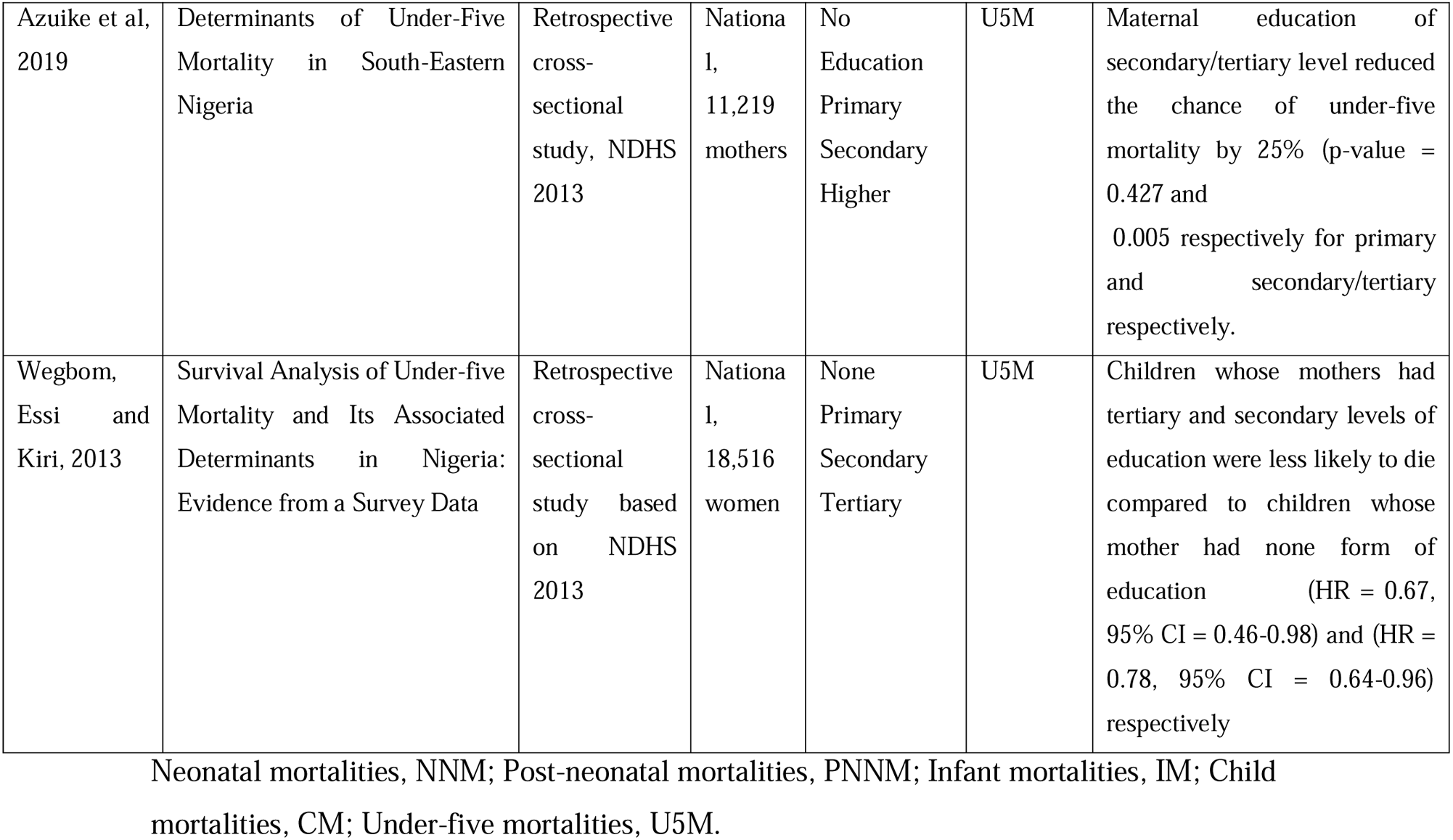
List of included studies and their characteristics.

**Figure 1:**
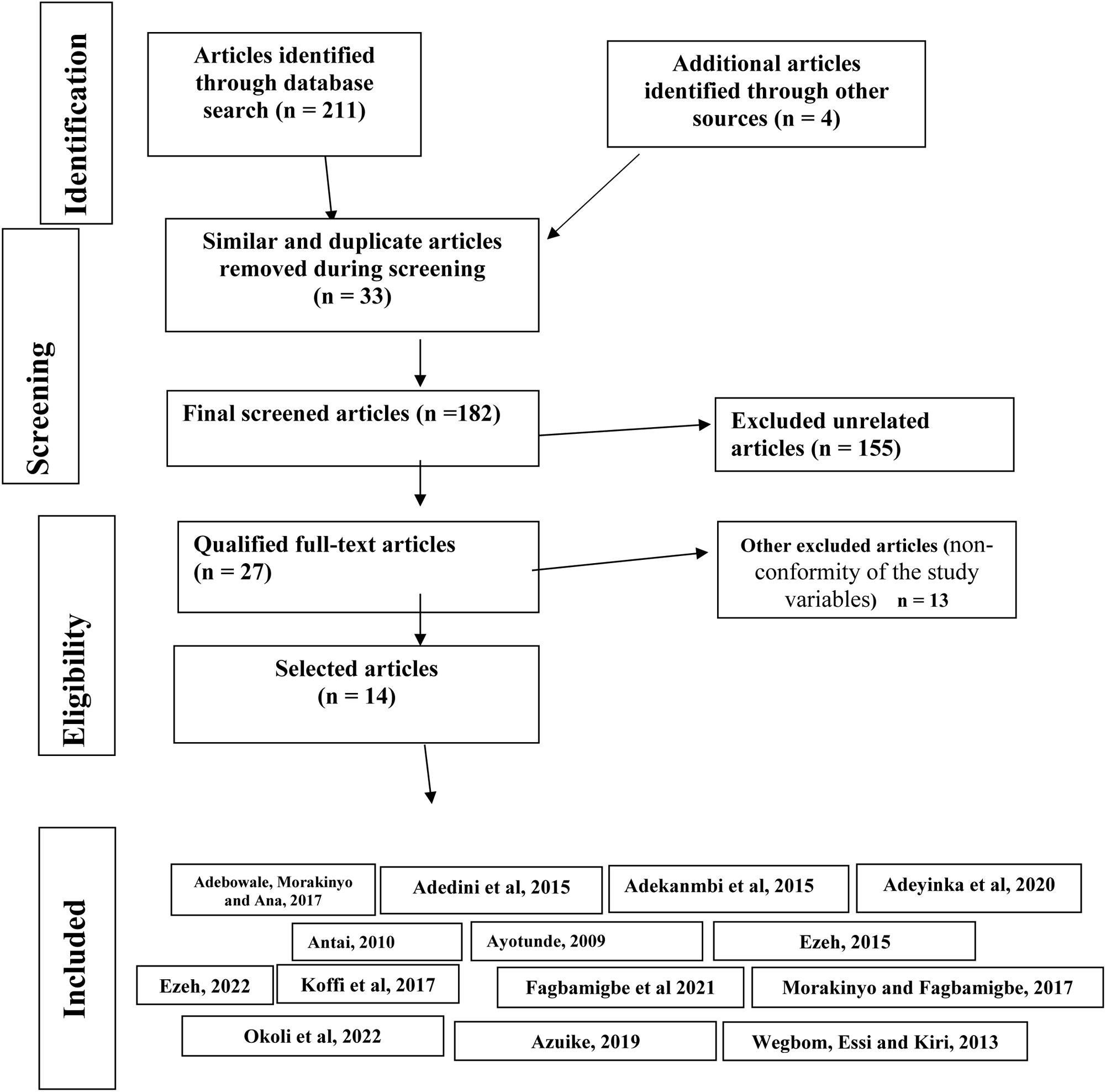
PRISMA flow chart demonstrating the steps for articles selection.

### Appraisal of included studies

Critical appraisal of the included studies was done using Critical Appraisal Tool for Analytical Cross-sectional Studies developed by the Joanna Briggs Institute (JBI)^15^. A second reviewer was used to appraise the data.

## Findings/ Results

A total of 215 articles were identified from the initial search of the databases and other sources. Out of this number, 33 were removed due to duplication. After screening the titles of the remaining 182, another 155 were excluded because their titles were not related to the topic of this systematic review. The abstracts of the remaining 27 articles were further screened and another 13 were further excluded due to non-conformity of the study variables with that of the index study. For example, measurement of IM rather than U5M. This gave rise to 14 remaining studies. However, some studies such as the one by Morakinyo and Fagbamigbe^16^, which measured NNM, IM and U5M were included while only the data on U5M was extracted and discussed. No studies were excluded based on methodical quality. Fourteen studies were therefore left for analysis and discussion. This is contained in figure 1.

### Study characteristics

All the included 14 evidence were retrospective cross-sectional studies based on the Nigeria’s DHS except the study by Adeyinka et al^17^ which utilized the nation’s Multiple Indicator Cluster Survey (MICS) of 2016/2017. Therefore, all included studies were based on secondary data (no primary research was found). The studies by Ezeh et al^18^ and Azuike ^19^ were sub-national while the remaining 12 were national.

Of the 13 NDHS-based studies, Antai^20^ and Ayotunde^21^ used the 2003 survey, Adedini et al^22^ used the 2008. Out of the remaining, Adebowale, Morakinyo and Ana,^23^ Adekanmbi et al^24^, Koffi et al^11^, Azuike et al^19^ and Wegbom, Essi and Kiri^25^ all used the 2013 survey for their studies at different times. In addition, Fagbamigbe et al^26^, and Okoli et al^27^ used the 2018 data. Finally, the remaining 3 used data from the 2003, 2008 and 2018 surveys for their report. These include Eze et al^28^, Morakinyo and Fagbamigbe^16^ and Ezeh et al^18^.

### Variables and their classification in the included studies

The outcome variable of interest was under-five mortality defined in this review as all child deaths after birth up till before the fifth birthday (0 to 59 months), while the independent variable was maternal education. Nine of the studies had U5M as an outcome variable, 2 had IM and U5M, one had NNM, IM and U5M, and one had PNNM, IM, CM and U5M, while another measured NNM, PNNM and U5M as outcome variables. However, for all the studies the outcome variable of interest in this SLR was U5M, while the selected studies had the independent variable (maternal education) similarly stratified into: None, Primary, Secondary and Higher/Tertiary education (Table 1).

### Critical appraisal of included studies

Critical appraisal summary of included studies is displayed on Table 2. All had relevant ethical approval while the objectives were clearly outlined and achieved using relevant methods. The sample sizes ranged from 7,620 to 79,953 and these were justified based on the set objectives. All authors clearly defined their sample frame and reference population while the non-response rate bias was all negligible. Appropriate validation tools were used to measure the variables and they were all analysed using appropriate tests of statistics. The different studies used different tests of statistical relationship such as adjusted odds ratio (aOR), hazard ratio (HR), p-values and incidence rate ratio (IRR), as appropriate. Some studies used descriptive statistics to display results, and these were considered appropriate based on set objectives. All the authors identified possible limitations to their studies, and these were considered adequate. In general, the included studies were rated of good quality and the risk for bias was considered minimal.

**Table 2:**
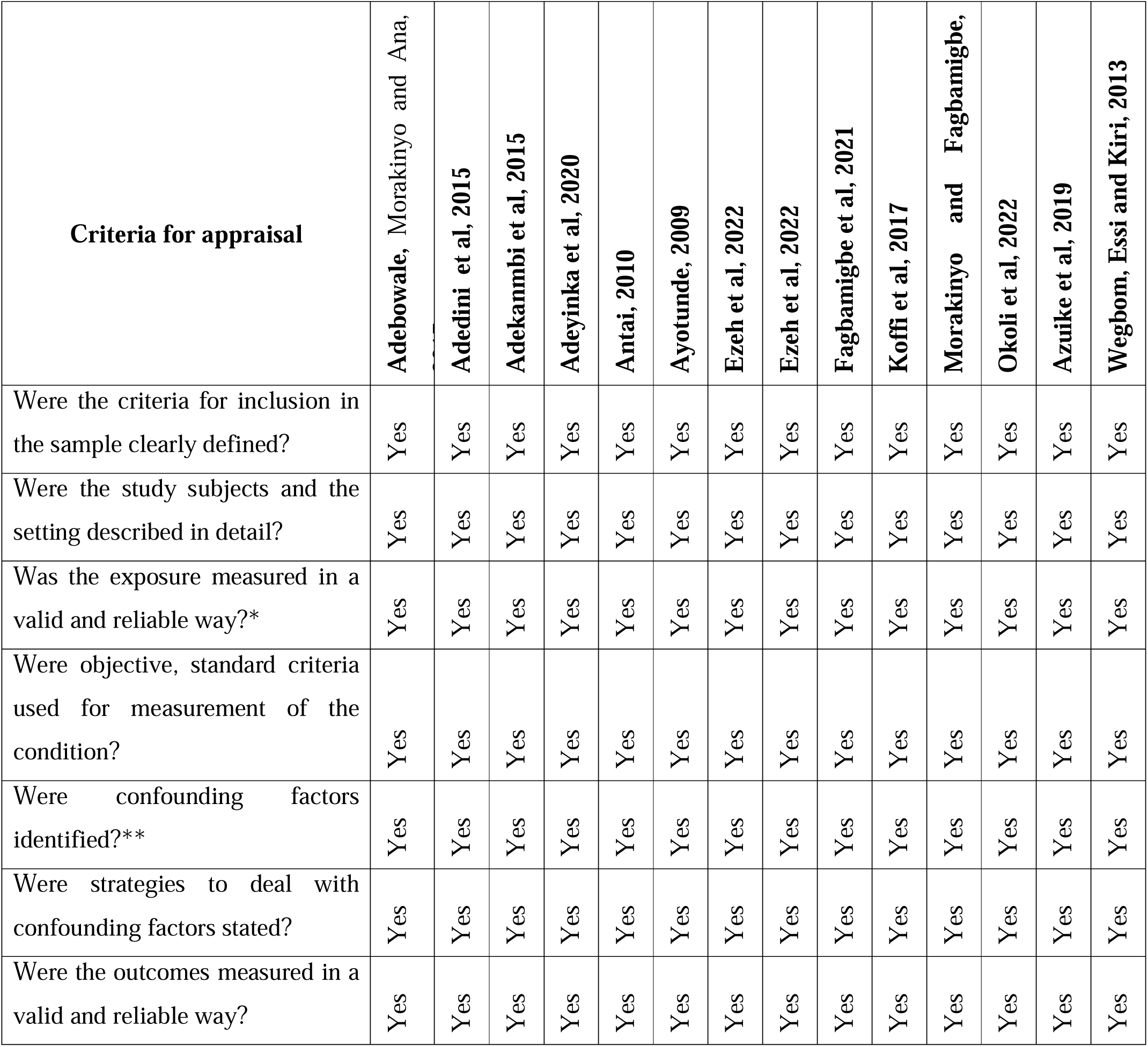
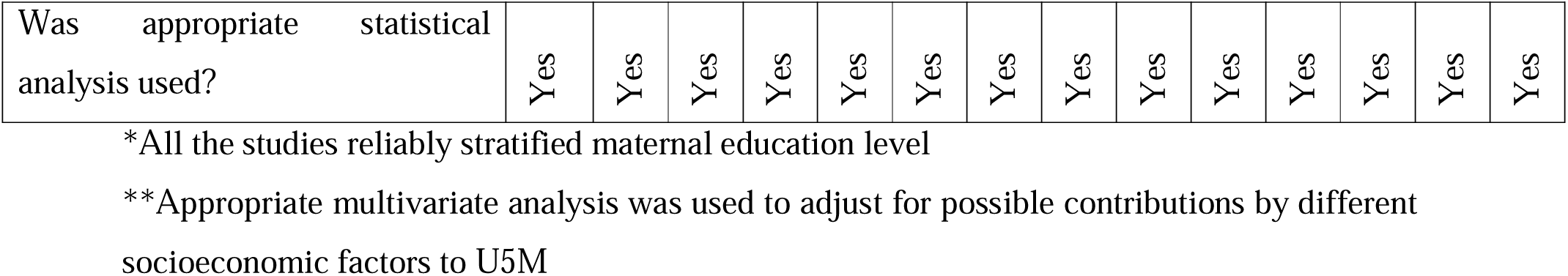
Critical appraisal summary.

### Major findings from the review

The result indicates that there was less odd of U5M among children whose mothers had at least primary education and further suggests that the risk of dying reduces further with higher levels of maternal education. According to Adedini et al^22^, there was a lower risk of dying among children whose mothers had secondary or higher education (HR: 0.84, p < 0.05), while Ezeh et al (2015) also agreed that children whose mothers had no formal education were more liable to dying (HR=1.19, CI 1.02 to 1.41). Similar to their findings, Wegbom, Essi and Kiri^25^ in their report expressed that children whose mothers had no formal education were more liable to dying (HR=1.19, CI 1.02 to 1.41), just as Fagbamigbe et al^26^ captured that there was a double risk of dying for children whose mothers had no form of education with an incidence rate ratio (IRR) of 2.14, at 95% credible interval (CrI) of 1.51 to 3.03. The findings by other authors were not different as Ezeh et al^18^ reported a significantly higher odd of dying for children whose mothers had no formal education (aOR = 1.53; 95% CI: 1.17– 2.01) when compared with children whose mothers had primary education (aOR = 1.80, 95% CI: 1.40–2.33). Others reported significant associations between maternal education and U5M. For example, Adebowale, Morakinyo and Ana,^23^ (p-value <0.001), Antai,^20^ (p-value 0.000), Ayotunde,^21^ (p-value 0.000), while Koffi et al^11^ reported that 72.5% of the uneducated women were from the northern part were majority of the U5M occurred (x^2^:723.8 p-value 0.000). In like manner, Adekanmbi et al^24^ concluded that Children whose mothers had secondary or higher education had higher odds of survival (p< 0.0001) while a descriptive analysis by Okoli et al^27^ suggested that U5M was 46.5% higher among children born to mothers with no formal education compared to their educated counterparts.

## Discussion

This SLR suggests that there is an association between maternal education and U5M and that children of mothers with higher levels of educational attainment are less prone to U5M compared to their counterparts with less educated mothers. For example, while comparing mothers without formal education with those who had at least primary education, Fagbamigbe et al^26^ concluded that the children of the former had a double risk of dying compared to the later. Whereas Adebowale, Morakinyo and Ana^23^ noted that U5M reduced with increasing level of maternal education, implying that there are hierarchies of U5M at different levels of maternal education. These findings have been corroborated by other researchers. For instance, Mandal, Paul and Chouhan^29^ reported from India that the level of maternal education significantly affected U5M in the region. Likewise, Bado, and Sathiya^30^ in 2016 observed that improved maternal education was partly responsible for achievements in the U5M figures within some selected sub-Saharan African countries. Consequently, maternal education is an inescapable knowledge gap to be addressed on the path to achieving U5M and the SDG 3.

This systematic review may be admired for the attributes of the selected studies. Firstly, all the studies were based on national survey data, the NDHS and the MICS. The NDHS are the largest data globally that provide child mortality data and other important indicators^31^. In addition, all but two of the studies had a national spread and that could further strengthen the validity of their conclusions. Furthermore, the use of multivariate regression analysis in most of them helped to reduce the contribution of other socio-economic cofounding variables. Likewise, no systematic review on the subject in Nigeria was cited, perhaps making this study novel. However, this SLR is not without limitations. To start with, some of the selected studies were based on same data sets. Therefore, the similar findings observed in them may not be surprising. Secondly, this study did not aim to conduct a meta-analysis, which may have added more strength to the conclusions. However, undertaking it may have been challenging due to the heterogeneity in analytical methods employed in the different studies. The high rate of under-five mortality in Nigeria has far reaching implication for globalization and global health. To exemplify, Nigeria is Africa’s most populous^32^, and according to the United Nations,^33^ the country is targeted to be the world’s 4^th^ most populated by 2050. It is therefore inferable that U5M in Nigeria could make direct and significant contributions to Africa’s, and indeed, global child mortality numbers. This could cause increased strain on limited global health resources.

Apart from maternal education, household wealth is another observed overarching risk factor for under five deaths in Nigeria^8^. According to Edeme, Ifelunini and Okereke^34^, the ability to procure health care services by individuals and households, which is determined by their level of financial and economic income, remains a major challenge to the attainment of quality standards in child survival in Nigeria. This factor, like others has a spiral effect and its consequences tend to ramify globally^8^. To stress the tie and mutual relevance of household income and globalization, Jai^35^ concluded that improvements in globalization deteriorated income inequalities in Korea. However, another scholar, though agreeing partly with this position, explained that the effect of globalization may not be one-sided^36^. The author opined that if complementary policies are in place, globalization may be gainful to the poor as well as wealthier populations. The author argued that macroeconomic stability could be promoted in different settings during globalization if complementary policies are institutionalized. Therefore, improvements in maternal education, household income and other conceptualized risk factors for U5M could directly improve the child health survival indices both locally and globally and even drive the attainment of the SDGs.

In addition to creating better access for female education, the SDGs’ attainment may have much to do with the performance of the MDGs. For example, despite the success recorded globally in the implementation of the MDGs, its performance in Nigeria was rated low as the country only managed to meet two out of the eight goals at the end of 2015^37^. According to the author, much is still needed to address critical goals such as child mortality, education, poverty reduction and environmental sustainability. It further pointed that the non-inclusion of players at the local level during implementation was one major reason for the poor performance of the MDGs, and therefore suggested that these factors should be priority while striving to achieve the SDGs. Meanwhile, according to the Danish Institute for Human Rights, as at 2020 (5 years into SDG implementation), the U5MR for Nigeria stood at almost 115 deaths per 1,000 live births^38^, almost five times over the 2030 target. This does not suggest a positive trajectory towards the attainment of the SDG3 in less than 10 years. However, all hope may not be lost according to Adesiyan^37^ who posited that the SDGs, and indeed SDG3, can be met if careful attention is given to the factors that mitigated the success of the MDG health goals. Those factors, according to the author, include the need for local ownership, improvement on data generation and monitoring, stronger local and international collaborations, more healthy partnership across the tiers of government as well as the need to explore other avenues for financial resources. In addition, the impact of the SDG3 depends on the success of other SDGs. For example, Davies et al recommends that for the SDG3 to be achieved at the end of its schedule, the government of Nigeria must address deficits in other sectors such as the challenges currently faced around power, fiscal corruption and infrastructural deficits in the country^39^. In addition, implementing the goal requires local adaptability. For example, Kenya is implementing the SDG3 selectively since the global goal aligns only partially with the country’s health priorities^40^. Therefore, on a global scale, the benefits of the SDGs will accrue for universal public health practice if countries identify their peculiar priorities and adapt the goals to suit these advantages and the global objective.

To address these issues, the role of global collaborative players like the world trade organization (WTO), the WHO, the World Bank, and international civil society organizations (CSOs) for health cannot be over-emphasized. For instance, to address issues related to fiscal and financial deficits, existing WTO and World Bank initiatives could come handy. Likewise, the WHO could serve to promote co-operation among scientific and professional groups which contribute to the advancement of health, provide appropriate technical assistance, and help to establish and maintain effective collaboration with the United Nations and other professional groups, among other responsibilities. These bodies working multilaterally in collaboration with the country directly and through CSOs could alleviate much of the factors that challenge the SDG’s progress and attainment.

## Conclusion

The SDGs are an universal call to action for countries to end poverty, protect the planet, and ensure that by 2030 all people enjoy peace and prosperity, while the SDG 3 focuses on ensuring healthy lives and promoting well-being for all at all ages. The reduction of U5M by 2030 is integral to the SDG3 and Nigeria has not fared well with the U5MR index over the years. In turn, maternal education is one of the fundamental socio-economic risk factors responsible for this drawback. And so, this systematic literature review hoped to find out from available evidence, the effect of maternal education on U5M and to as well determine how gaps in maternal education could impact the attainment of the SDG target for U5M and the SDG3, in the country.

To summarize, 14 retrospective cross-sectional studies were reviewed, and the result suggest that there was less odd of U5M among children whose mothers had at least primary education. The studies further revealed that the risk of dying reduced further with higher maternal educational level. In addition, it suggested that maternal education is an inescapable factor and a knowledge gap to be addressed on the path to achieving SDG3.

The systematic review had the strength of being based primarily on a majority national surveys with sound study techniques that had little risk of study bias. It may however be faulted on the ground that several of the included studies used the same data sets thereby making their corroborative findings non-surprising.

## Data Availability

All data produced in the present work are contained in the manuscript.

## Acknowledgements

The author acknowledges the authors whose publications were cited and reviewed in this work.

## References

1. Kumar S, Kumar N, Vivekadhish S. Millennium Development Goals (MDGs) to Sustainable Development Goals (SDGs): Addressing Unfinished Agenda and Strengthening Sustainable Development and Partnership. Indian Journal of Community Medicine. 2016;41(1):1–4. doi: 10.4103/0970-0218.170955.

2. United Nation Development Programme (2020) Sustainable Development Goals. Available at: https://www.undp.org/content/undp/en/home/sustainable-development-goals.html/ (Accessed 20 August 2022).

3. Lucas AO, Gilles HM. Short textbook of public health medicine for the tropics. 4th edn. London: Hodder Arnold. 2003:17–20.

4. Pedersen J, Liu J. Child mortality estimation: appropriate time periods for child mortality estimates from full birth histories. PLoS Medicine 2012; 9(8): e1001289. doi:10.1371/journal.pmed.1001289.

5. UNICEF Data, Great Britain. Monitoring the situation of children and women. Available at: https://data.unicef.org/country/gbr/. (Accessed 5th August 2022).

6. UNICEF Data, Nigeria. Monitoring the situation of children and women. Available at: https://data.unicef.org/country/nga/. (Accessed 5th August 2022).

7. Sharma RK. Causal pathways to infant mortality: linking social variables to Infant mortality through intermediate variables. Journal of Health and Social Policy. 1998; 9(3):15–28. 10.1300/J045v09n03_02/

8. Van Malderen C, Amouzou A, Barros AJ, Masquelier B, Van Oyen H, Speybroeck N. Socioeconomic factors contributing to under-five mortality in sub-Saharan Africa: a decomposition analysis. BMC public health. 2019 Dec;19:1–9.

9. Shifa GT, Ahmed AA, Yalew AW. Socioeconomic and environmental determinants of under-five mortality in Gamo Gofa Zone, Southern Ethiopia: a matched case control study. BMC international health and human rights. 2018 Dec;18:1–1.

10. Kanmiki, E.W., et al. (2014) ‘Socio-economic and demographic determinants of under-five mortality in rural northern Ghana’, BMC International Health and Human Rights 14(24) 10.1186/1472-698X-14-24.

11. Koffi AK, Kalter HD, Loveth EN, Quinley J, Monehin J, Black RE. Beyond causes of death: The social determinants of mortality among children aged 1-59 months in Nigeria from 2009 to 2013. PLoS one. 2017 May 31;12(5):e0177025.

12. Smith-Greenaway, E. (2013) ‘Maternal reading skills and child mortality in Nigeria: a reassessment of why education matters’, Demography, 50(5), pp.1551–1561. DOI 10.1007/s13524-013-0209-1.

13. Liberati A, Altman DG, Tetzlaff J, Mulrow C, Gøtzsche PC, Ioannidis JP, Clarke M, Devereaux PJ, Kleijnen J, Moher D. The PRISMA statement for reporting systematic reviews and meta-analyses of studies that evaluate health care interventions: explanation and elaboration. Annals of internal medicine. 2009 Aug 18;151(4):W–65.

14. Kiross GT, Chojenta C, Barker D, Tiruye TY, Loxton D. The effect of maternal education on infant mortality in Ethiopia: A systematic review and meta-analysis. PloS one. 2019 Jul 29;14(7):e0220076.

15. The Joanna Briggs Institute Critical Appraisal Tools for Use in JBI Systematic Reviews (2019) Checklist for Analytical Cross Sectional Studies. Available at: https://jbi.global/sites/default/files/2019-05/JBI_Critical_Appraisal-Checklist_for_Analytical_Cross_Sectional_Studies2017_0.pdf/ (Accesses 9 August 2022).

16. Morakinyo OM, Fagbamigbe AF. Neonatal, infant and under-five mortalities in Nigeria: an examination of trends and drivers (2003-2013). PLoS One. 2017 Aug 9;12(8):e0182990.

17. Adeyinka DA, Muhajarine N, Petrucka P, Isaac EW. Inequities in child survival in Nigerian communities during the Sustainable Development Goal era: insights from analysis of 2016/2017 Multiple Indicator Cluster Survey. BMC Public Health. 2020 Dec;20:1–8.

18. Ezeh OK, Odumegwu AO, Oforkansi GH, Abada UD, Ogbo FA, Goson PC, et al. Trends and factors associated with under-5 mortality in northwest Nigeria (2008– 2018). Annals of Global Health. 2022;88(1).

19. Azuike EC, Onyemachi PE, Amah CC, Okafor KC, Anene JO, Enwonwu KG, et al. Determinants of under-five mortality in South-Eastern Nigeria. J Community Med Public Health Care. 2019;6:049.

20. Antai, D. Inequalities in under-5 mortality in Nigeria: do ethnicity and socioeconomic position matter? Journal of Epidemiology. 2011;21(1):13–20. 10.2188/jea.JE20100049.

21. Ayotunde T, Mary O, Melvin AO, Faniyi FF. Maternal age at birth and under-5 mortality in Nigeria. East African Journal of Public Health. 2009 Apr 1;6(1):11–4.

22. Adedini SA, Odimegwu C, Imasiku EN, Ononokpono DN, Ibisomi L. Regional variations in infant and child mortality in Nigeria: a multilevel analysis. Journal of biosocial science. 2015 Mar;47(2):165–87.

23. Adebowale SA, Morakinyo OM, Ana GR. Housing materials as predictors of under-five mortality in Nigeria: evidence from 2013 demographic and health survey. BMC pediatrics. 2017 Dec;17:1–3.

24. Adekanmbi VT, Kandala NB, Stranges S, Uthman OA. Contextual socioeconomic factors associated with childhood mortality in Nigeria: a multilevel analysis. J Epidemiol Community Health. 2015 Nov 1;69(11):1102–8.

25. Wegbom AI, Essi ID, Kiri VA. Survival analysis of under-five mortality and its associated determinants in Nigeria: evidence from a survey data. International Journal of Statistics and Applications. 2019;9(2):59–66.

26. Fagbamigbe AF, Salawu MM, Abatan SM, Ajumobi O. Approximation of the Cox survival regression model by MCMC Bayesian Hierarchical Poisson modelling of factors associated with childhood mortality in Nigeria. Scientific reports. 2021 Jun 29;11(1):13497.

27. Okoli CI, Hajizadeh M, Rahman MM, Khanam R. Geographic and socioeconomic inequalities in the survival of children under-five in Nigeria. Scientific Reports. 2022 May 19;12(1):8389.

28. Ezeh OK, Agho KE, Dibley MJ, Hall JJ, Page AN. Risk factors for postneonatal, infant, child and under-5 mortality in Nigeria: a pooled cross-sectional analysis. BMJ open. 2015 Mar 1;5(3):e006779.

29. Mandal S, Paul P, Chouhan P. Impact of maternal education on under-five mortality of children in India: insights from the National Family Health Survey, 2005–2006 and 2015–2016. Death studies. 2021 Nov 26;45(10):788–94.

30. Bado AR, Sathiya Susuman A. Women’s education and health inequalities in under-five mortality in selected sub-Saharan African countries, 1990–2015. Plos one. 2016 Jul 21;11(7):e0159186.

31. Rutstein SO. Factors associated with trends in infant and child mortality in developing countries during the 1990s’, Bullettin of the World Health Organization. 2000 78: 1256–1270 Available at: https://www.ncbi.nlm.nih.gov/pmc/articles/PMC2560619/pdf/11100620.pdf/ (Acceseed 18 August 2022).

32. Reed HE, Mberu BU. Capitalizing on Nigeria’s demographic dividend: reaping the benefits and diminishing the burdens. Etude de la population africaine=African population studies. 2014 Mar;27(2):319.

33. United Nations. World Population Prospects: The 2012 Revision. Population Division of the Department of Economic and Social Affairs of the United Nations Secretariat. Available at: https://www.un.org/en/development/desa/publications/world-population-prospects-the-2012-revision.html/ (Accessed 22 August 2022).

34. Edeme RK, Ifelunini IA, Okereke OS. Relationship between household income and child mortality in Nigeria. Am J Life Sci. 2014;2:1–2.

35. Mah JS. The impact of globalization on income distribution: the Korean experience. Applied Economics Letters. 2002 Dec 1;9(15):1007–9.

36. Harrison, A. Globalization and Poverty 2006, 12347. NBER Working Paper Series. National Bureau of Economic Research 1050 Massachusetts Avenue Cambridge, MA 02138 June 2006. Available at: https://www.nber.org/system/files/working_papers/w12347/w12347.pdf/ (Accessed August 25 2022).

37. Adesiyan EA. Local government and the attainment of sustainable development goals in Nigeria: lessons from the millennium development goals’, African Journal of Governance and Development, 2018;7(1):50–62. Available at: https://journals.co.za/doi/epdf/10.10520/EJC-f6567ef4f/ (Accessed 18 August 2022).

38. The Danish Institute for Human Rights. The Human Rights Guide to the Sustainable Development Goals 2015. Available at: https://sdg.humanrights.dk/en/goals-and-targets/ (Accessed 8 August 2022).

39. Davies IE, Nwankwo CO, Olofinnade OM, Michaels TA. Insight review on impact of infrastructural development in driving the SDGs in developing nations: A case study of Nigeria. InIOP Conference Series: Materials Science and Engineering 2019 Nov 1 (Vol. 640, No. 1, p. 012112). IOP Publishing. Available at https://iopscience.iop.org/article/10.1088/1757-899X/640/1/012112/pdf/ (Accessed 18 July 2024).

40. Kamau E, MacNaughton G. The impact of SDG 3 on health priorities in Kenya. Journal of Developing Societies. 2019 Dec;35(4):458–80.

